# Assessing the Hospital Surge Capacity of the Kenyan Health System in the Face of the COVID-19 Pandemic

**DOI:** 10.1101/2020.04.08.20057984

**Authors:** Edwine Barasa, Paul Ouma, Emelda Okiro

## Abstract

**Introduction:** The COVID-19 pandemic will test the capacity of health systems worldwide. Health systems will need surge capacity to absorb acute increases in caseload due to the pandemic. We assessed the capacity of the Kenyan health system to absorb surges in the number of people that will need hospitalization and critical care because of the COVID-19.

**Methods:** We assumed that 2% of the Kenyan population get symptomatic infection by SARS-Cov-2 based on modelled estimates for Kenya and determined the health system surge capacity for COVID-19 under three transmission curve scenarios, 6, 12, and 18 months. We estimated four measures of hospital surge capacity namely: 1) hospital bed surge capacity 2) ICU bed surge capacity 3) Hospital bed tipping point, and 5) ICU bed tipping point. We computed this nationally and for all the 47 county governments.

**Results:** The capacity of Kenyan hospitals to absorb increases in caseload due to COVID-19 is constrained by the availability of oxygen, with only 58% of hospital beds in hospitals with oxygen supply. There is substantial variation in hospital bed surge capacity across counties. For example, under the 6 months transmission scenario, the percentage of available general hospital beds that would be taken up by COVID-19 cases varied from 12% Tharaka Nithi county, to 145% in Trans Nzoia county. Kenya faces substantial gaps in ICU beds and ventilator capacity. Only 22 out of the 47 counties have at least 1 ICU unit. Kenya will need an additional 1,511 ICU beds and 1,609 ventilators (6 months transmission curve) to 374 ICU beds and 472 ventilators (18 months transmission curve) to absorb caseloads due to COVID-19.

**Conclusion:** Significant gaps exist in Kenya’s capacity for hospitals to accommodate a potential surge in caseload due to COVID-19. Alongside efforts to slow and supress the transmission of the infection, the Kenyan government will need to implement adaptive measures and additional investments to expand the hospital surge capacity for COVID-19. Additional investments will however need to be strategically prioritized to focus on strengthening essential services first, such as oxygen availability before higher cost investments such as ICU beds and ventilators.

## INTRODUCTION

A cluster of pneumonia cases of unknown origin were reported in Wuhan, Hubei province in China in December 2019 (1). The causative microbe was later identified as the novel enveloped RNA beta corona virus named Severe acute respiratory syndrome coronavirus 2 (SARS-CoV-2), and the disease named COVID-19 (2). The ensuing COVID-19 outbreak has spread from China to over 209 countries and territories worldwide, infecting over 1.2 million individuals and killing over 60,000 people as at 6^th^ April 2020 (3). COVID-19 has since been declared a public health emergency of international concern (PHEIC) and a pandemic by the World Health Organization (4).

Epidemics present a challenge to health systems because they cause an acute increase in the demand for health services (5,6). A critical aspect of country response to an epidemic is hence the health system surge capacity; that is the health system’s capacity to absorb and accommodate the acute increase in the demand of healthcare services (7). Unfortunately health systems are typically designed for average healthcare demand rather than epidemics (7). Health system surge capacity for COVID-19 entails both hard and soft elements. Hard elements include infrastructure, health workforce, and healthcare commodities, while soft elements include response coordination, logistics for needed supplies, protocols and guidelines for prevention mitigation and containment, and effective communication (7,8).

COVID -19 is an acute respiratory infection placing demands for the health system to screen and test suspected cases, contact tracing of exposed individuals and isolation of cases, and severe cases requiring hospitalization, with critical cases requiring intensive care (9–11). Countries that have experienced sudden surges in COVID-19 cases such China, Iran, Italy, Spain and the United States, some with more sophisticated and well-resourced health systems, show evidence of overwhelmed health systems with hospitals unable to accommodate the sudden surge in the number of patients needing hospitalization and critical care (12,13).

Estimating the surge capacity of the health system in the face of epidemics provides useful information for planning, mobilization and allocation for response (5,6). Kenya reported its first case on the 13^th^ of March 2020. Since then, the number of confirmed cases have been steadily increasing and modelling estimates predict that about 2% of the Kenya population could be infected with SARS-Cov-2 and develop symptoms (symptomatic infected) (14). In this paper, we estimate the surge capacity of the Kenyan health system in the face of the COVID-19 pandemic. Specifically, we assess the capacity of the Kenyan health system to absorb a surge in the number of people that will need hospitalization and critical care as a direct result of COVID-19. We also estimate the health system tipping points, that is the maximum number of symptomatic infected cases that the Kenyan health system can absorb given the current capacity and make recommendations on measures that could be adopted to enhance the resilience of the Kenyan health system to the COVID-19 pandemic.

## METHODS

### Data Sources

To map out capacity of hospitals to provide COVID-19 services, we used data from the master health facility list (MFL), the harmonised health facility assessment (HHFA) and a survey from the Kenya health federation on number of ICU beds and ventilators. Data on number of beds was obtained from the MFL and the HHFA. Those with oxygen capacity were mapped from recent work aimed at mapping hospital services in Kenya and involved triangulation of data from HHFA, the district health information system and the 2013 service availability and readiness assessment. ICUs and ventilators were mapped from the HHFA and a nationwide survey commissioned by the Kenya Health Federation to assess the capacity of hospitals to manage COVID-19 patients. This data is provided in Supplemental Table 1.

**Table 1:** Health system capacity for COVID 19 measures

| Measure | Definition |
| --- | --- |
| (1) General hospital bed surge capacity | The proportion of available general hospital beds with oxygen supply, in both the private and public sector in Kenya, that will be required to care for COVID-19 patients. |
| (2) ICU bed surge capacity | The proportion of available ICU beds (with ventilators), in both the private and public sector in Kenya, that will be required to care for COVID-19 patients |
| (3) General hospital bed tipping point | The maximum number of SARS-Cov-2 symptomatic infections, whose expected number of hospitalizations can be absorbed by the available number of general hospital beds with oxygen in both the public and private sector in Kenya |
| (4) ICU bed tipping point | The maximum number of SARS-Cov-2 symptomatic infections, whose expected number of ICU admissions can be absorbed by the available number of ICU beds with oxygen in both the public and private sector in Kenya |

### Estimating General Hospital, and ICU bed Surge Capacity

We define surge capacity as the available capacity to accommodate healthcare demand arising from an epidemic (7,15). This is in addition to existing capacity for non-epidemic healthcare demands. We computed four measures of health system hospital surge capacity for COVID-19 nationally and by county, namely; 1) general hospital bed surge capacity 2) ICU bed surge capacity, 3) health system general hospital bed tipping point, and 5) health system ICU bed tipping point. Table 1 outlines the definitions of each of the capacity measures. We determined the health system surge capacity for COVID-19 under three transmission curve scenarios, 6, 12, and 18 months. A 6 months transmission curve means that all the expected number of symptomatic infections will occur within 6 months resulting in a higher peak, compared to when the same number of symptomatic infections are spread out over 18 months.

A hospital bed has been defined as one that is staffed and maintained for continuous medical care for inpatients in hospitals (16). In this analysis we only count hospital beds in hospitals that have oxygen supply because oxygen is an essential requirement for the management of COVID-19 patients. We do not include neonatal cots or day care beds in our count. An ICU on the other hand has been defined as a separate area of a hospital that is specially equipped and staffed to manage and monitor patients with life-threatening conditions that include organ system failure (such as respiratory, cardiovascular, renal system support) (17). In this analysis we counted ICU beds that have accompanying ventilators since ventilators are required to provide mechanical ventilation for COVID-19 patients that are critically ill. We did not find data on the functionality of these ICUs and whether they meet the norms of an ICU with regards to staffing, equipment other than ventilators.

First, we computed the number of available general hospital beds (*H*_*a*_) by multiplying the total number of general hospital beds (*H*_*t*_) by the complement of the average general hospital bed occupancy rate (*O*_*h*_) (equation 1). We used the same approach to compute the number of available ICU beds (*ICU*_*a*_) that are available (equation 2), where *ICU*_*t*_ is the total number of ICU beds in Kenya (public and private), and *O*_*i*_ is the average ICU bed occupancy rate.

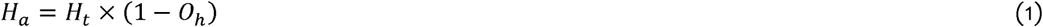

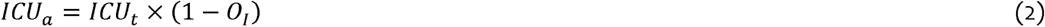

Second, we determined the number of people that are likely to need hospital admission (*C*_*h*_), and critical care (*C*_*i*_) by multiplying the age specific risks of severe (*R*_*s*_) and critical (*R*_*c*_) disease by the number of people that are likely to get symptomatic infection in the Kenyan population (*P*_*i*_) (equation 4 and 5). We assumed that approximately 2% of the population will be infected by SARs-Cov-2 and develop symptoms based on model estimates for Kenya (14) and applied this to the Kenyan population. We obtained the age specific risks of severe and critical disease from a published analysis of 2,449 cases in the United States (11).

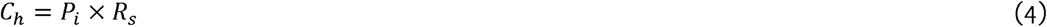

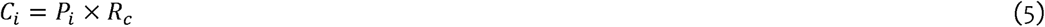

Third, we determined the number of general hospital bed days (*H*_*bd*_) that would be needed by COVID-19 patients by multiplying the number of patients needing hospital admission by the expected average length of stay (ALOS) (equation 6). We used the same approach to determine the number of ICU bed days (*ICU*_*bd*_) that would be needed by COVID-19 patients (equation 7). We assumed that the average length of stay for patients admitted with COVID-19 is 12 days (10).

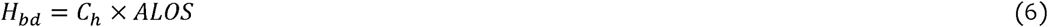

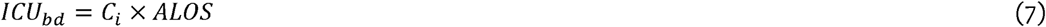

Fourth, to determine the proportion of available general hospital beds that would be utilized (health system surge capacity measure 1) (*H*_*sc*_) under each of the transmission curve scenarios, we divided the number of hospital bed days by the multiple of available hospital beds and the scenario duration in days (*d*) (either 6, 12, or 18 months) (equation 8). We applied the same approach to compute the proportion of available ICU beds that would be utilized (health system surge capacity measure 2) (*ICU*_*sc*_) (equation 9).

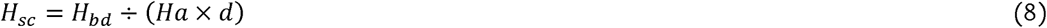

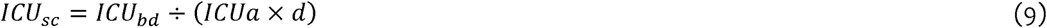

Lastly, we determined hospital tipping point (health system surge capacity measure 3) (*H*_*t*_) by dividing the number of available hospital bed days under each transmission curve scenario by the risk of hospitalization (equation 10). We applied the same approach to compute the ICU bed tipping point (health system surge capacity measure 4) (*ICU*_*t*_).

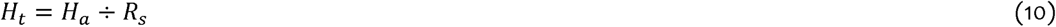

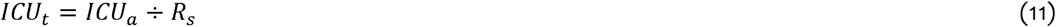

### Mapping Geographical Access to ICU units

We mapped geographical access to ICU units because they are a critical constraint. Proximity to hospitals with available ICUs was computed using a cost distance algorithm described in detail elsewhere (18). In brief a cost friction surface that accounts for both walking and motorised transport was developed using different travel time speeds across land uses, road networks and elevation, while also correcting for availability of barriers such as protected areas and water bodies. This surface was then used to compute travel time to the nearest ICU hospital at 100m spatial resolution for the whole country. County level quotients for population within 2 hours to the nearest hospital were then extracted by overlaying the travel time surface to a gridded population layer at 100m spatial resolution. The model was implemented with the assumption of cross border movement across the different counties.

## FINDINGS

### Hospital bed surge capacity

While Kenya has 64,181 hospital beds across all sectors (public, faith based/NGO, private for profit), only 37,216 (58%) of these beds are in hospitals that have oxygen supply. Figure 1 outlines findings on hospital bed surge capacity nationally under different transmission curve scenarios. The findings reveal that when the number of general hospitals beds nationally are considered, projected COVID-19 cases needing hospitalization will take up 39%, 19%, and 13% of available hospital beds if the pandemic lasts 6, 12, and 18 months respectively.

**Figure 1:**
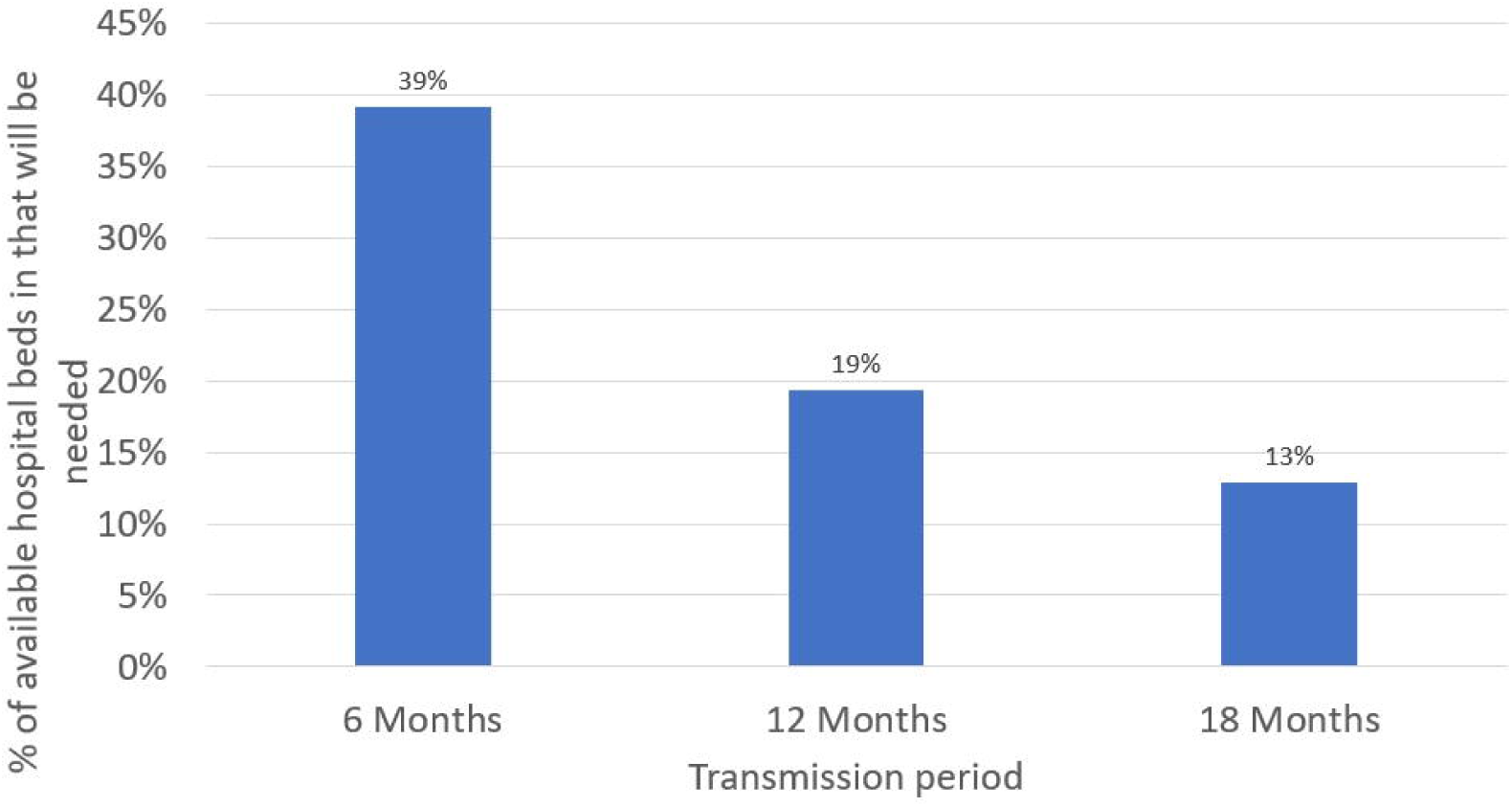
Proportion of available hospital beds that would be required if 2% of the population are infected and develop symptoms

However, hospital bed surge capacity varies across the 47 counties. For example, under the 6 months transmission scenario, the percentage of available general hospital beds that would be taken up by COVID-19 cases varied from 12% Tharaka Nithi county, to 145% in Trans Nzoia county. Figure 2 shows maps of hospital surge capacity for the 47 counties under the three different transmission scenarios.

**Figure 2:**
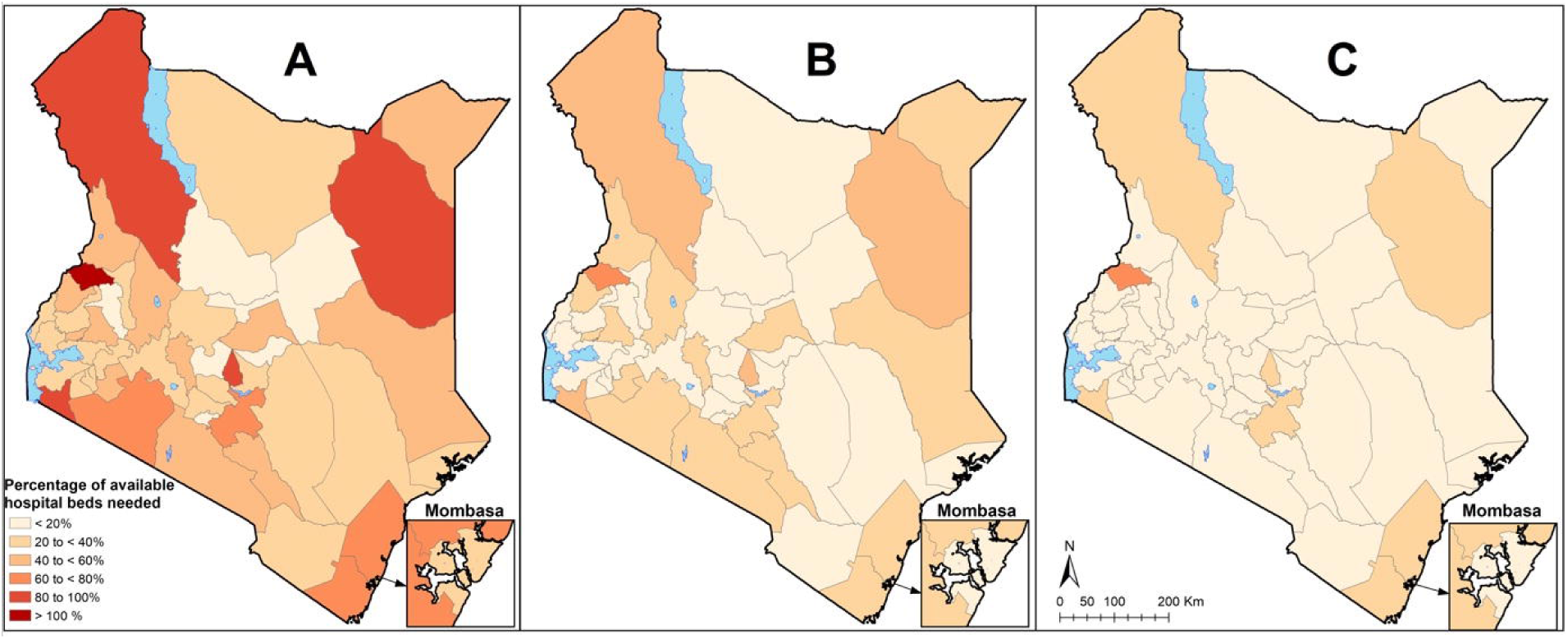
General hospital bed surge capacity for the 47 counties under the three different transmission scenarios. The maps show % of available general hospital bed capacity that will be needed if 2% of the population are infected and develop symptoms over under A) the 6-month transmission scenario, B) the 12-month transmission scenario and C) the 18-month transmission scenario. Proportions are shown to increase from light red to dark red.

### ICU bed Surge Capacity

While Kenya has 537 ICU beds, it only has 256 ventilators. Therefore, when ventilators are considered, 281 of existing ICU beds do not have the accompanying equipment to provide care for COVID-19 critically ill patients. Figure 3 outlines findings on ICU bed surge capacity nationally under different transmission curve scenarios. The findings reveal that if the pandemic is concentrated over 6 months, Kenya will on average need 1,511 additional ICU beds to absorb COVID-19 cases, and an additional 1,609 ventilators. If the transmission curve is flattened to 12 months, or 18 months, Kenya will need on average 650 or 374, of additional ICU beds respectively, and 748 and 472 ventilators respectively.

**Figure 3:**
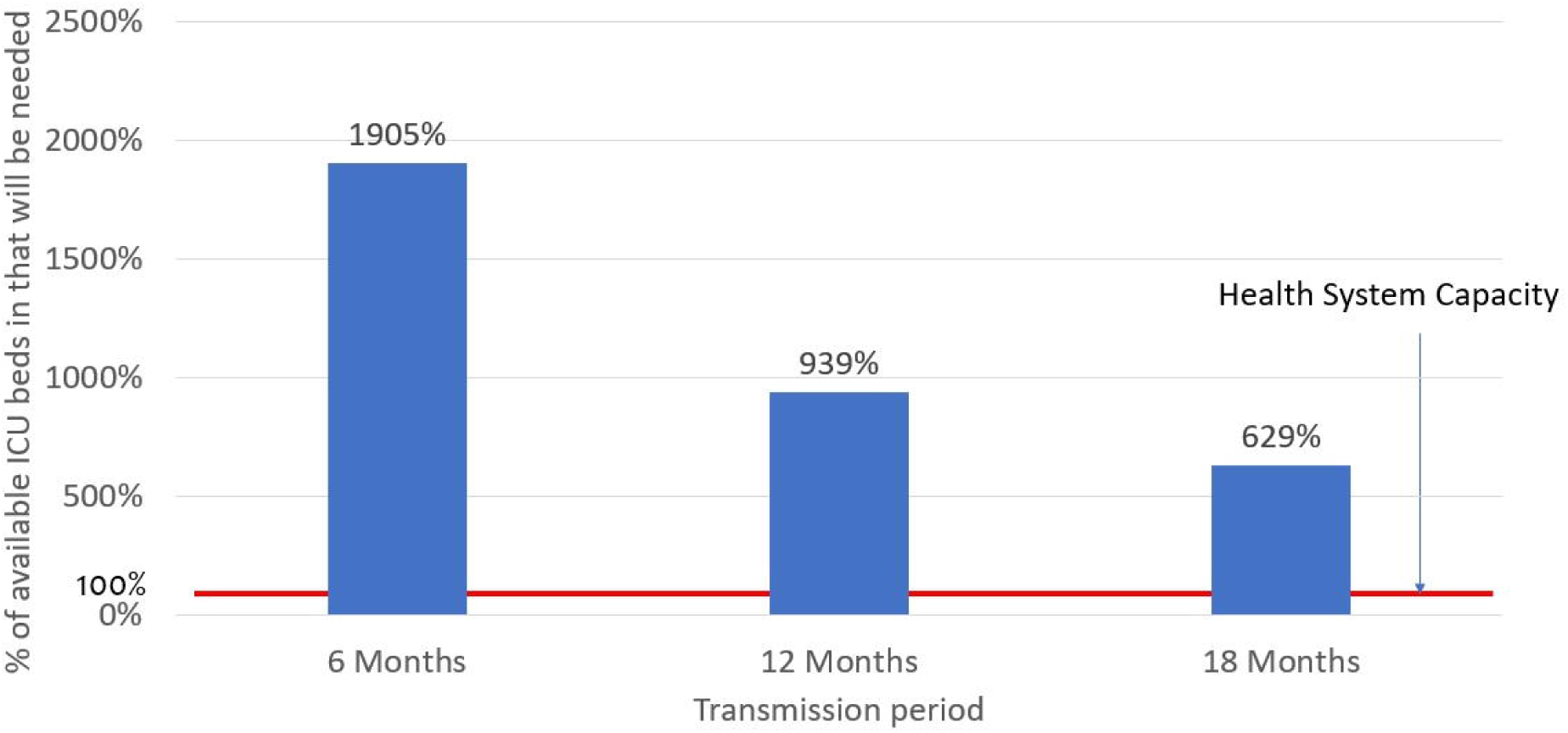
Proportion of available ICU beds that would be required if 2% of the population are infected and develop symptoms

Only 22 out of the 47 counties have at least 1 ICU unit signalling significant geographical disparities. When availability was considered, only seven counties were found to have at least one ICU bed available.

### Geographical Access to ICU units

Access to the existing ICU units is also significantly constrained. Only 22% of Kenya’s population lives within 2 hours of a facility with an ICU available, with only Vihiga county having more than 80% of its population living within this time threshold. Other counties with relatively good accessibility quotients are Bungoma, Kisumu, Kiambu and Nairobi with more than 60% currently living within a facility having an ICU available. In 25 counties, no one lives within 2 hours of a facility that has an available ICU. Figure 4 shows variations the percentage of the population within 2 hours of the nearest ICU hospital across the 47 counties in Kenya.

**Figure 4:**
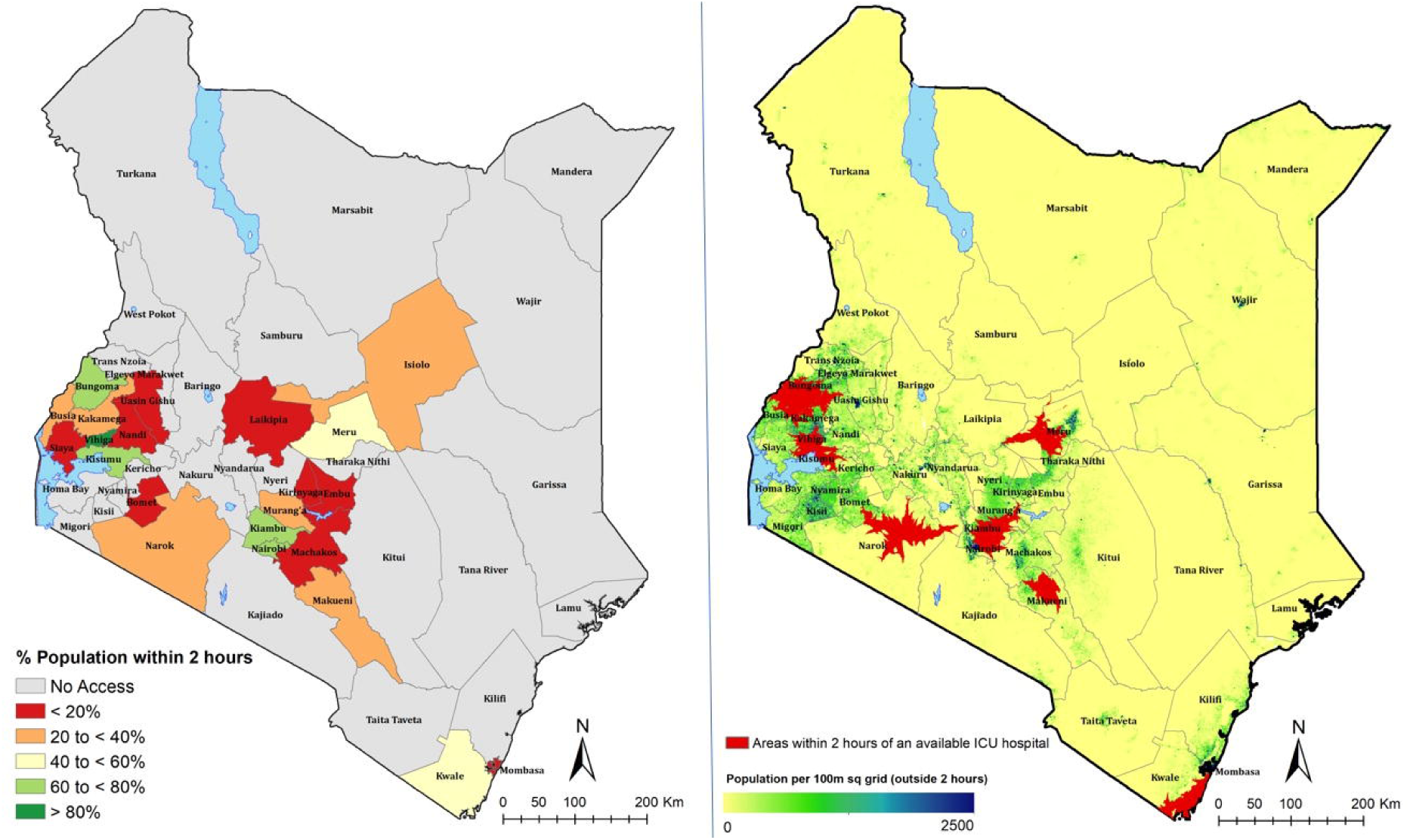
A) % population within 2 hours of the nearest available ICU hospital, with increasing accessibility from red to dark green. Counties with no access are shown in grey. B) Map showing populated places outside the 2 hour travel time.

### Health system general hospital bed and ICU bed tipping points

Table 2 outlines the general hospital bed, ICU bed, and ventilator health system tipping points nationally under the different transmission curve scenarios. General hospital bed tipping point varies from 2.8 million number of symptomatic infections if the pandemic is concentrated over 6 months to 8.5 million symptomatic infections if the pandemic is spread over 18 months. On the other hand, ICU bed and ventilators tipping points varies from 121,137 number of symptomatic infections for ICU beds, and 86,156 number of symptomatic infections for ventilator if the pandemic is concentrated over 6 months to 366,777 symptomatic infections for ICU beds and 260,862 for ventilators if the pandemic is spread over 18 months.

**Table 2:** Hospital Bed and ICU bed tipping points

| Transmission Curve Scenario | Maximum number of symptomatic that can be accommodated by existing hospital bed capacity | Maximum number of symptomatic that can be accommodated by existing ICU bed capacity |
| --- | --- | --- |
| 6 months | 2,809,221 | 57,749 |
| 12 months | 5,696,475 | 117,102 |
| 18 months | 8,505,695.65 | 174,851 |

There is variation in the maximum number of clinical cases that can be absorbed by the general hospital bed and ICU bed capacity across counties. For instance, the general hospital bed tipping point ranged from 11,532 clinical cases in Lamu county to 560,038 clinical cases in Nairobi county, while ICU bed tipping point varied from zero symptomatic cases in 38 counties to 43,116 symptomatic cases in Nairobi county under the 6 month transmission curve scenario. Figure 5 & 6 presents variations in tipping points across the 47 counties under each of the 3 transmission curve scenarios for hospitals and ICU beds respectively.

**Figure 5:**
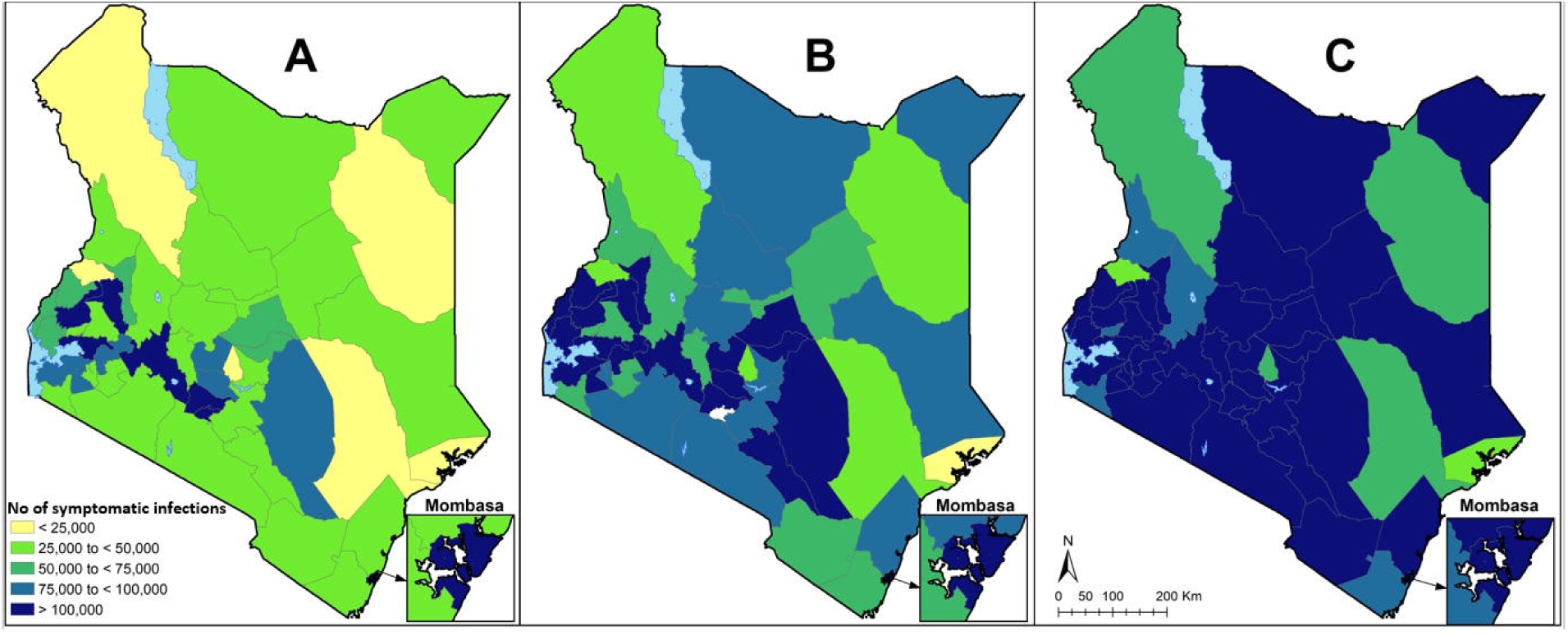
Number of COVID-19 symptomatic infections that can be absorbed by current hospital bed capacity over A) 6 months, B) 12 months and C) 18 months

**Figure 6:**
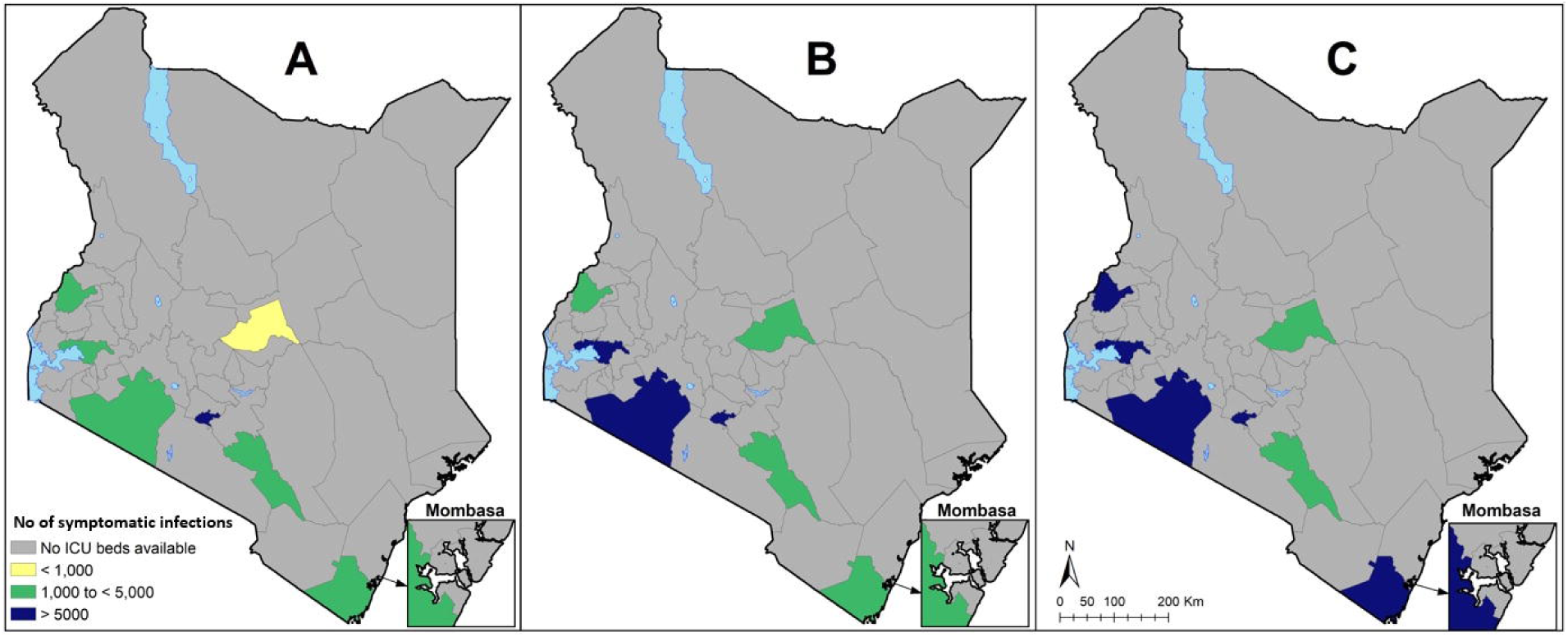
Number of COVID-19 symptomatic infections that can be absorbed by current ICU bed capacity over A) 6 months, B) 12 months and C) 18 months.

## DISCUSSION

This analysis quantifies the gaps in general hospital bed, ICU bed, and ventilator capacity to absorb the shock of the COVID-19 pandemic in Kenya and highlights the geographical variation in capacity across the 47 Kenyan counties. The analysis finds that while Kenya has no surge capacity for critical care under the 3 transmission scenario’s, there may be some surge capacity for general hospital beds, if other aspects of capacity are discounted. However, hospital beds are an inadequate proxy of hospital capacity and give only a partial view of health system capacity. Rather, it would be ideal to assess capacity in terms of, among others, a comprehensive package of required infrastructure, trained healthcare workers, medical supplies, and governance/institutional arrangements (7). For instance, while fully functional ICU beds are crucial, they are not useful in the absence of an adequate number of trained critical care health workers and/or medical supplies that are required for case management. In the context of COVID-19, capacity to test, and to protect healthcare workers with personal protective equipment (PPE) have been identified as crucial. General gaps in human resources and essential medicines in Kenya have been documented (19). Unfortunately, granular data on these dimensions of health system capacity was not available, and even the data available are of questionable specificity. For instance, while we obtained data on ICU beds nationally and at county level, we could not obtain data on the functionality and equipment of these ICU’s and hence whether they are fit for purpose. Therefore, the capacity gaps we report are underestimated and it is likely that when all other inputs are considered, the capacity of the Kenyan health system to absorb increased caseloads due to COVID-19 will be significantly lower. For instance, while hospital beds may be available to take additional patients with COVID-19, the existing health workforce shortages cannot accommodate this. Similar challenges have been faced before in Africa. For instance, the 2014-2016 Ebola epidemic overwhelmed the health systems of West African countries (20–23).

Our scenarios assume a uniform proportion of symptomatic infections across counties, while, like the experience of other countries has shown, there are likely to be hotspots within the country that will account for a disproportionate amount of COVID-19 cases. In the more realistic scenario where any or several counties account for a disproportionate proportion of infected cases in Kenya, the health system will come under even more pressure. It is also important to highlight that while our scenarios have assumed that 2% of the Kenyan population will get symptomatic COVID-19 infection, this is a modelled estimate (14) and the true value could be much lower or higher. It is also not clear how the pandemic will affect individuals and systems in Africa given that Africa not only has a different (much younger) population structure, but also has other vulnerabilities that include malnutrition, HIV/AIDs, Tuberculosis, Malaria, alongside an emerging burden of non-communicable diseases.

These limitations notwithstanding, our findings provide an indication of the challenges that the Kenyan health system will face if the COVID-19 epidemic progresses unabated. The findings emphasize the analysis in other settings that slowing the rate of spread of the infection could minimize the demands on the healthcare system (24). It also highlights the need for strengthening health system surge capacity through both adaptive measures and additional investments in needed capacities. From the experience of other countries, adaptive measures that can be explored to expand hospital bed capacity include a) postponing elective procedures, b) re-assessing patients that are currently admitted and discharging those that can be safely discharged and managed at home and c) refurbishing and transitioning existing hospital spaces into to makeshift ward areas and provision of oxygen. The Kenyan government is already undertaking some of these measures and is also considering non-hospital spaces. For instance, the government has earmarked educational institutions (high schools, colleges, universities) with boarding facilities as potential isolation centres. The government will also need to collaborate with the private sector and leverage on healthcare resources in the private sector. Our analysis assumed that both public and private sector hospital beds and ICUs will be available which may require certain arrangements to be put in place. Given that the private sector accounts for about 50% of healthcare facilities in Kenya, restricting the response to the public sector will reduce the health system capacity by approximately 50%.

In addition to these adaptive measures, the government will also need to make urgent additional investments to expand health system capacity. In doing so however, the government will need to be pragmatic and prioritize these investments in terms of what to invest in and when to optimize impact, given existing system gaps and financial constraints. For instance, it is not practical to invest in additional ICU beds in the short term, given the substantial gaps that currently exist, the massive resources required to do so, and the companion capacity gaps of skilled critical care health workers that require long term training to get into the system. Also, investing in ICU care assumes that general hospital care for patients that do not require critical care and ventilators is adequate. In practice, only 37,216 hospital beds out of 64,181 hospital beds in Kenya are in healthcare facilities that have oxygen supply. This reduces the number of hospital beds that can provide care to COVID-19 patient with severe disease by 42%. Further, a recent health facility assessment found that only 16% of healthcare facilities in Kenya have pulse oximeters, a vital device for monitoring oxygen saturation and therapy (19). This survey further found that the mean availability of tracer items for emergency breathing interventions (pulse oximeters, micronebulizer, beclomethasone and salbutamol inhalers, oxygen with tubing, flowmeter, and humidifier, resuscitation bags, intubation devices with connecting tube, chest tubes with insertion sets, and CPAP equipment) was only 13% (19). It may be more pragmatic for Kenya, and other LMICs in the same situation to first invest in making existing hospital beds functional by providing oxygen and procuring and making these devices available before focusing on ICU beds and ventilators. This is not only more financially and operationally feasible but will save more lives given that the share of COVID-19 patients with severe disease, and requiring inpatient admission with oxygen is about 5 times greater than those likely to have critical disease needing ICU care. Once basic hospital care capacity has been improved, it might make sense then to improve the functionality of existing critical care facilities before setting up additional ones. For instance, our findings show that while Kenya has 537 ICU beds, it only has 256 ventilators, rendering more than half the existing ICUs impractical in the face of COVID-19. Equipping and staffing existing ICUs might be a priority in the medium to long term. Further, while we did not assess health workforce capacity gaps, previous analyses have shown serious health workforce gaps in Kenya that will undoubtedly affect the country’s COVID-19 response (19). Given the risk of infection of health workers (25), the first measure should be to protect the existing health workers from getting sick so that they can continue to provide needed care. This is especially important considering reports of PPE shortages. Additionally, in a context where a substantial number of core health workers that include nurses and doctors are unemployed, the government should hire these currently unemployed health workers even if on a temporary basis to deal with the current shortfall. In allocating resources to increase the health system capacity, the government should be guided by likely differences in burden of the epidemic but also the differences in capacity gaps across the 47 counties. Optimal resource allocation guided by need will ensure that resources are used optimally and equitably and enhance the overall capacity of the health system.

Finally, our analysis highlights the potential value of granular data on availability and location of health services to inform community response and the need to make this information publicly available. Yet, most countries in Africa do not have readily available and accessible inventories of coverage and distribution of facilities nor do they have comprehensive assessments of capacity and services available within these facilities (26). In addition, high resolution data on population density, movement patterns and understanding where vulnerable populations such as informal settlement are located provide critical additional data layers to support the epidemic response. Such data make it increasingly possible to carry out a more contextualized analysis that reflects the complexity of vulnerability, allow better prediction of disease spread and refine demand and supply forecasting.

Currently, data on infrastructure, equipment and human resource remains fragmented, and we had to rely on multiple sources of data, including a survey commissioned as a result of the COVID-19 outbreak to map available services in hospitals. We were unable to obtain adequate number of health workforce available at each facility, further emphasizing the challenges of data collection. With the country having a robust master health facility list that is regularly updated, further investment should be directed towards updating the services available, as this can be useful in adequately allocating resources for expansion of services.

## Data Availability

The data used for this analysis has been provided as supplementary information

## ACKNOWLEDGMENTS

This paper is submitted with the permission of the director, KEMRI. Edwine Barasa is supported by the Wellcome Trust core award grant to KEMRI-Wellcome Trust Research Programme (number 203077). Paul Ouma is supported by the DELTAS grant awarded to IDeAL/KEMRI-Wellcome Trust Research Programme (number 107769). Emelda Okirois supported by a Wellcome Trust Intermediary Fellowship (number 201866). We would like to Acknowledge Prof Robert Snow, Prof Philip Bejon, and Dr. Ambrose Agweyu for comments and feedback on earlier versions of the manuscript. We would also like to acknowledge Dr. Elizabeth Amakove Wala of the Kenya Healthcare federation for facilitating data access from the Kenya Healthcare Federation.

